# Motivation and concerns regarding the use of preprints among Brazilian health researchers: a survey

**DOI:** 10.64898/2026.09.11.26362857

**Authors:** Alessandra Pertile de Oliveira, Lara Dotto, Mayara Colpo Prado, Rafaela Oliveira Pilecco, Bernardo Antonio Agostini, Gabriel Kalil Rocha Pereira, Rafael Sarkis-Onofre

**Author notes:** **Corresponding author** Rafael Sarkis-Onofre.

## Abstract

This study aimed to evaluate the motivations and concerns related to preprint use among Brazilian health researchers. A mixed-methods online survey of all 1,596 holders of CNPq Research Productivity Grants in seven health fields was conducted in 2025. The questionnaire was adapted from a survey of bioRxiv authors, translated into Portuguese, and pilot-tested. Closed-ended responses were analyzed descriptively, and free-text responses were grouped by similarity. Overall, 178 researchers accessed the survey (11.2%), and 143 answered items on preprints and were included in the final analysis (9.0% effective response rate). The mean age was 55.9 years, and 88.0% were affiliated with public institutions. Regarding corresponding-author publications over the previous five years, 62.2% of respondents had deposited no preprints, while 37.8% had deposited at least one. Among preprints users, 79.2% agreed that deposition was freely decided by the authors. The main motivations were rapid sharing of findings (69.8%) and obtaining feedback (49.1%). Few reported benefits in terms of citations (17.3%) or online dissemination (16.9%). Among those who had not deposited preprints, the main reason was not wanting to deposit them (57.6%), followed by being unaware of the option (35.3%). Of the 22 free-text responses, 19 reflected an informed decision not to deposit, most commonly because peer review was considered necessary before public dissemination. Preprint use was limited mainly by a lack of intention to deposit rather than a lack of awareness. Faster dissemination and feedback were the main motivations for using preprints.

## INTRODUCTION

The term “open science” refers to a set of actions and tools aimed at making scientific processes more transparent across all stages of research, from conception to the dissemination of results and research data (Lewis, 2020). The open science movement represents a new way of conducting science, based on collaborative and transparent strategies developed in response to limited access, insufficient reporting in scientific articles, and concerns about misconduct in many scientific studies (Lewis, 2020).

The practices promoted by open science enable data sharing, provide unrestricted access to knowledge, ensure transparency of information, and help researchers examine the implications of their findings, facilitating the identification of research with questionable practices (Vazire, 2017). Overall, these efforts and strategies aim to advance scientific knowledge and promote good research practices. Allen and Mehler (2019) emphasized that adopting open science methods enhances research credibility, although challenges remain regarding researchers’ motivation and adoption.

Among open science tools, preprints have received increasing attention because they allow authors to openly share original manuscripts on dedicated servers before formal peer review. By using this tool, authors can establish credit for their work on a specific topic through a unique identifier, such as a DOI. In addition, preprints can increase the dissemination and visibility of scientific discoveries, enabling broader feedback from other researchers, who may identify flaws or errors or contribute data that strengthens the arguments presented in the study. Despite these potential advantages, preprints remain among the least adopted open science practices in the health and biomedical sciences, exhibiting marked heterogeneity across disciplines, career stages, and geographical regions (Pennington et al., 2026).

Understanding the motivations and concerns related to the use of preprints can help governments and funding agencies promote better research ecosystems by implementing appropriate policies and incentives to overcome barriers and foster a more open, collaborative, and efficient scientific environment, ultimately benefiting society. Brazil provides an important context for examining this issue because it combines substantial scientific output with persistent constraints in research funding, making it a relevant setting for understanding how open science practices are adopted in resource-limited research systems. Thus, this study aimed to evaluate the motivations and concerns related to preprint use among Brazilian health researchers.

## METHODOLOGY

The present study was designed as a mixed-methods survey study, including both quantitative and qualitative components. The article is reported based on the recommendations of the Checklist for Reporting of Survey Studies guidelines (Sharma et al., 2021).

### Protocol and Ethical aspects

The study protocol was registered on the Open Science Framework platform (https://osf.io/futch/overview). In addition, all study materials, including the questionnaire, are available on the platform.

The project was submitted to the local ethics committee and was approved under review number 6.930.655. All eligible participants were informed about the study objectives, risks, and benefits associated with the procedures, and those who agreed to participate signed the informed consent form.

### Sample

The target population comprised Brazilian health researchers awarded Research Productivity Grants (PQ) by the National Council for Scientific and Technological Development (CNPq), the main federal agency supporting scientific research in Brazil. This grant is awarded to distinguished researchers across different areas of knowledge and is intended to support and recognize researchers with established scientific contributions. The Research Productivity Grant is currently classified into three levels: level C, corresponding to the entry category; level B, corresponding to the development category; and level A, corresponding to researchers with fully consolidated research activity. In the national classification system adopted by Brazilian higher education and funding agencies (CNPq and CAPES), the Health Sciences domain encompasses seven disciplines: Dentistry, Medicine, Public Health, Nursing, Pharmacy, Physical Education, and Nutrition. This domain was selected because debates regarding rapid dissemination through preprints and the necessity of prior peer review are particularly relevant in health research, as findings can directly impact public health.

Using the publicly available <u>CNPq database</u>, we retrieved the complete registry of active productivity fellows across the seven health disciplines in July 2023. A total of 1,596 eligible researchers were identified, and all were invited to participate, thereby establishing a nationwide census approach to minimize sampling bias.

### Procedures and Questionnaire

Initially, 1,596 scholarship holders registered in July 2023 were identified. Their email addresses were collected from the Lattes Curriculum platform (a national information system managed by the Brazilian government), recently published articles, or the authors’ institutional webpages.

After email collection, researchers received an invitation to answer the questionnaire through the SurveyMonkey platform. Since the survey was anonymous and did not include a respondent identification field, all eligible researchers received two reminder emails: the first two weeks after the initial invitation and the second one month after the first reminder. The survey was conducted between March and April 2025.

The questionnaire consisted of two blocks. The first block included questions related to the researcher’s area of activity, scholarship level, and type of institution with which the researcher was affiliated. The second block included questions related to motivations for using or not using preprints, considering articles for which the researcher was the corresponding author and articles for which the researcher was not the corresponding author. For the purposes of this study, the following concept was adopted:

- Preprints: preliminary reports of studies that have not been certified by the peer review process (medRxiv, n.d.).

The questionnaire used in this study was based on a previously developed questionnaire that evaluated the use of preprints by corresponding authors of preprints posted on the bioRxiv platform (Fraser et al., 2022). It was translated into Portuguese by two researchers and reviewed by a third, all of whom were Brazilian and proficient in English. Prior to the start of the study, the translated version of the questionnaire was pilot tested with ten researchers affiliated with graduate programs in the areas covered by the study.

The two blocks of the questionnaire comprised a total of 23 questions - mostly closed-ended, with some open-ended questions included. No question in the questionnaire was mandatory; therefore, respondents could leave any question unanswered and proceed to the next one. The estimated time to complete the questionnaire was up to 15 minutes. The complete questionnaire is available on the Open Science Framework platform.

### Data analysis

Data were analyzed descriptively. Categorical variables were summarized using absolute and relative frequencies, while age was reported as a mean. Responses to the five-point Likert-scale items were summarized according to each response category and presented in tables and bar charts. Because no questionnaire item was mandatory, analyses were based on the available responses, and the denominator for each variable corresponded to the number of participants who answered that item. Free-text responses were reviewed and grouped into categories based on similarities in content, and the frequency of responses within each category was reported. No inferential analyses or statistical hypothesis tests were performed.

## RESULTS

A total of 1,596 researchers were invited to participate in the survey. Of these, 178 accessed the questionnaire and answered at least one question (11.2% overall response rate). Analyses were restricted to the 143 respondents who answered at least one item on preprints, yielding a final response rate of 9.0%.

Table 1 presents the sociodemographic characteristics of the respondents. The mean age was 55.9 years, and gender distribution was relatively balanced, with a slight male majority (54.2%). The most frequent fields among Research Productivity Grant holders were Dentistry (27.3%) and Medicine (26.6%). Regarding grant level, level C was the most frequent category (41.8%), and the vast majority of respondents were affiliated with public institutions (88.0%). Regarding publication volume as corresponding author in the previous five years, 47.6% of respondents reported publishing 21 or more articles. Over this same period, most respondents (62.2%) reported that none of their corresponding-author articles had been deposited as preprints, while 37.8% reported depositing at least some articles. No respondent reported that all articles had been deposited as preprints. Percentages are calculated on the respondents who answered each item, and the denominator for each characteristic is reported in Table 1.

Among respondents who reported having deposited some of their articles as preprints, the decision to deposit was made freely by the authors, with 79.2% agreeing or strongly agreeing (Fig. 1). In contrast, institutional open science policies did not appear to play a major role in this decision, as 32.1% of respondents strongly disagreed and 35.8% disagreed that depositing articles as preprints was necessary to comply with institutional open science policies. A similar pattern was observed for funding agency policies, with 39.6% strongly disagreeing and 41.5% disagreeing. When respondents were asked whether depositing their articles as preprints was their own suggestion, 22.6% agreed and 17.0% strongly agreed. However, disagreement with this statement was also frequent, indicating heterogeneity in how the decision to post preprints was initiated among researchers. In contrast, co-authors did not appear to be a major source of influence in the decision to post preprints: when asked whether depositing articles as preprints was usually suggested by their co-authors, 11.5% of respondents strongly disagreed, 32.7% disagreed, and 26.9% were neutral.

**Fig. 1.**
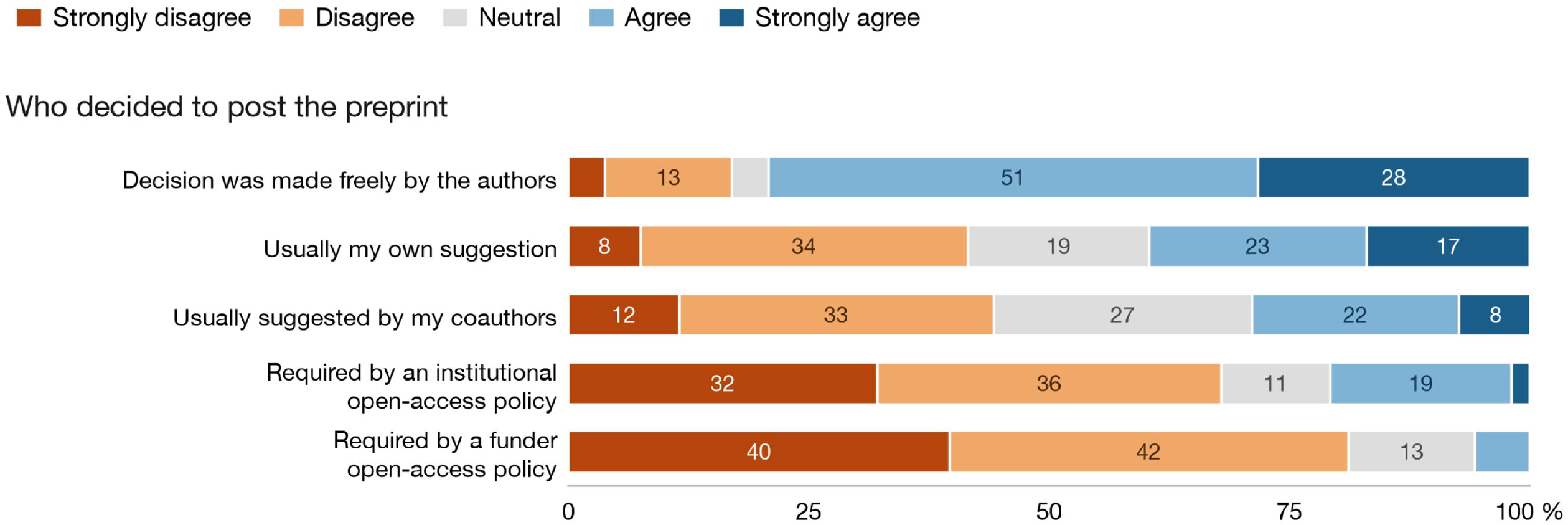
Level of agreement with statements about how the decision to post a preprint was made, among corresponding authors who posted at least some of their articles as preprints (n = 52–53 per statement). Numbers inside the bars are percentages of respondents; segments below 7% are left unlabelled

In terms of motivations for depositing preprints (Fig. 2a), sharing findings more rapidly received the most support, with 69.8% of respondents agreeing or strongly agreeing. It was followed by receiving more feedback on their work (49.1%), increasing awareness of their research (45.3%), and claiming priority for their findings (43.3%). The idea that preprints benefit the scientific enterprise received the least support, with 11.3% agreeing or strongly agreeing and 60.4% disagreeing or strongly disagreeing. Perceived benefits of posting preprints on article impact showed a similar pattern across the evaluated items (Fig. 2b). Overall, respondents tended to be neutral or to disagree that preprint deposition had benefits in terms of citations or other forms of online dissemination.

**Fig. 2.**
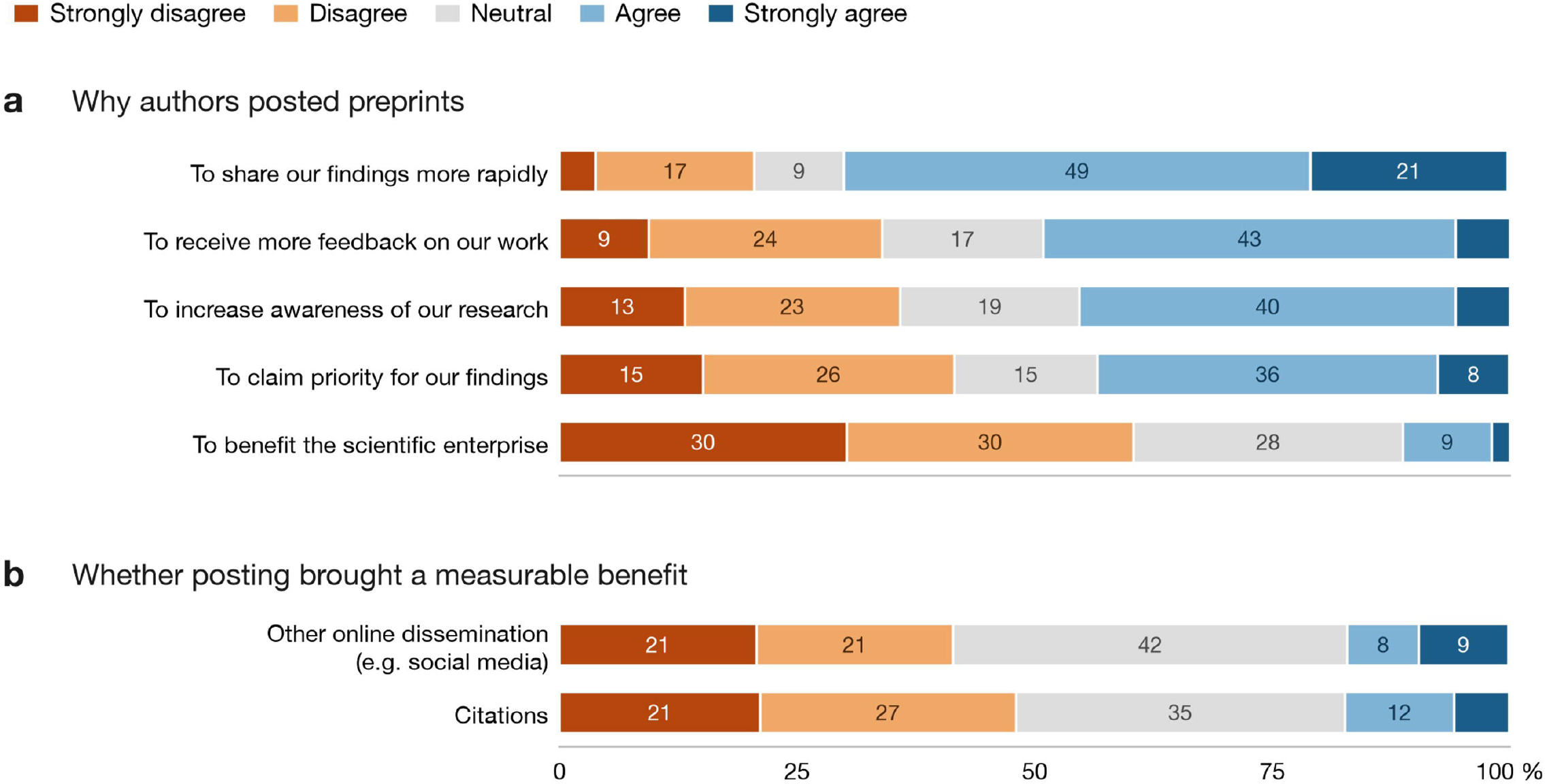
Motivations for posting preprints (a) and perceived benefits of having posted them (b), among corresponding authors who posted at least some of their articles as preprints (n = 52–53 per statement). Numbers inside the bars are percentages of respondents; segments below 7% are left unlabelled

Among non-adopters, reasons for not depositing preprints varied across domains (Fig. 3). Lack of awareness showed mixed responses: although 38.8% strongly disagreed that they were unaware of preprint options (indicating prior knowledge), 27.1% agreed with it. For the statement that the journals to which respondents intended to submit did not allow prior dissemination as preprints, the most frequent response was neutral (42.3%).

**Fig. 3.**
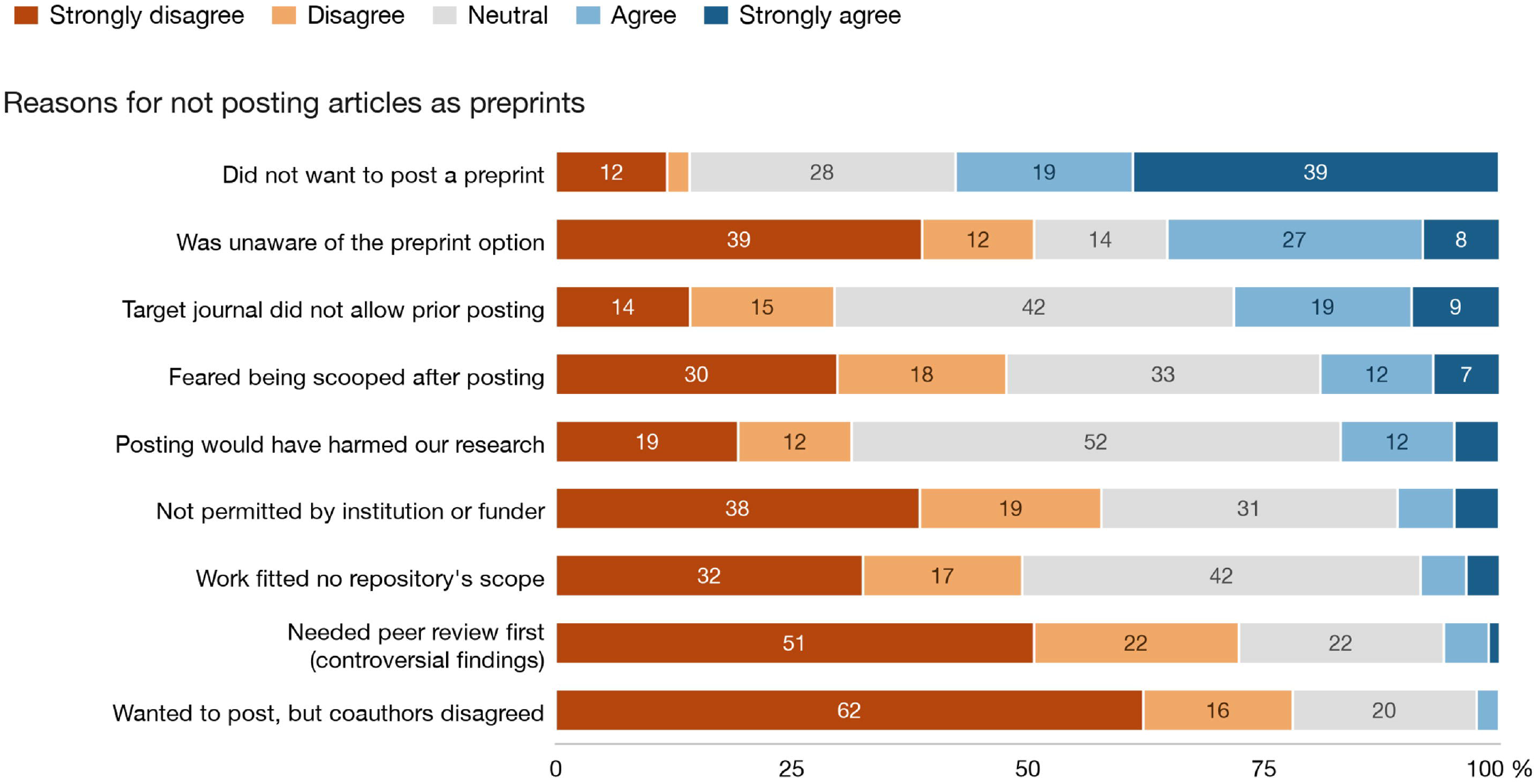
Level of agreement with possible reasons for not posting preprints, among corresponding authors who reported posting none of their articles as preprints (n = 82–85 per statement). Numbers inside the bars are percentages of respondents; segments below 7% are left unlabelled

Neither institutional nor funder restrictions were reported as major barriers to preprint usage. Instead, the primary barrier was a deliberate lack of intention: 57.6% of non-adopters agreed or strongly agreed (38.8% strongly agree, 18.8% agree) that they simply did not wish to deposit their manuscripts as preprints, while 28.2% remained neutral. Similarly, lack of fit with the scope of preprint repositories did not appear to be a major reason, as responses were concentrated in the neutral, disagree, and strongly disagree categories: 42.2% were neutral, 16.9% disagreed, and 32.5% strongly disagreed.

Twenty-three participants provided free-text responses explaining their reasons for not depositing preprints, of which 22 were informative (Supplementary Material). Content analysis yielded seven categories grouped under two main themes. Nineteen responses described an informed decision not to deposit the article as a preprint, meaning that the respondent was aware of the option but chose not to do so. Within this theme, the most frequent category was a preference for peer review before public dissemination, treated as a requirement rather than as a matter of utility (8 responses), followed by the absence of any perceived added value (5), the perception that deposition would jeopardize subsequent journal publication or attract predatory solicitation (3), the additional workload and lack of habit (2), and the absence of recognition of preprints in academic evaluation (1). The remaining three answers described non-use in the absence of an informed position: two respondents reported not knowing about the option or not having looked for information about it, and one reported having no formed opinion about preprints.

Among respondents who reported depositing some articles as preprints, perceptions about differences between articles posted as preprints and those not posted as preprints were generally mixed (Fig. 4). No clear pattern was observed for scientific quality, novelty, social value, journal impact factor, or expected citation impact. The only notable finding was related to online dissemination: 42.6% of respondents agreed that articles deposited as preprints were expected to be disseminated more widely online, for example on social media, than articles not deposited as preprints.

**Fig. 4.**
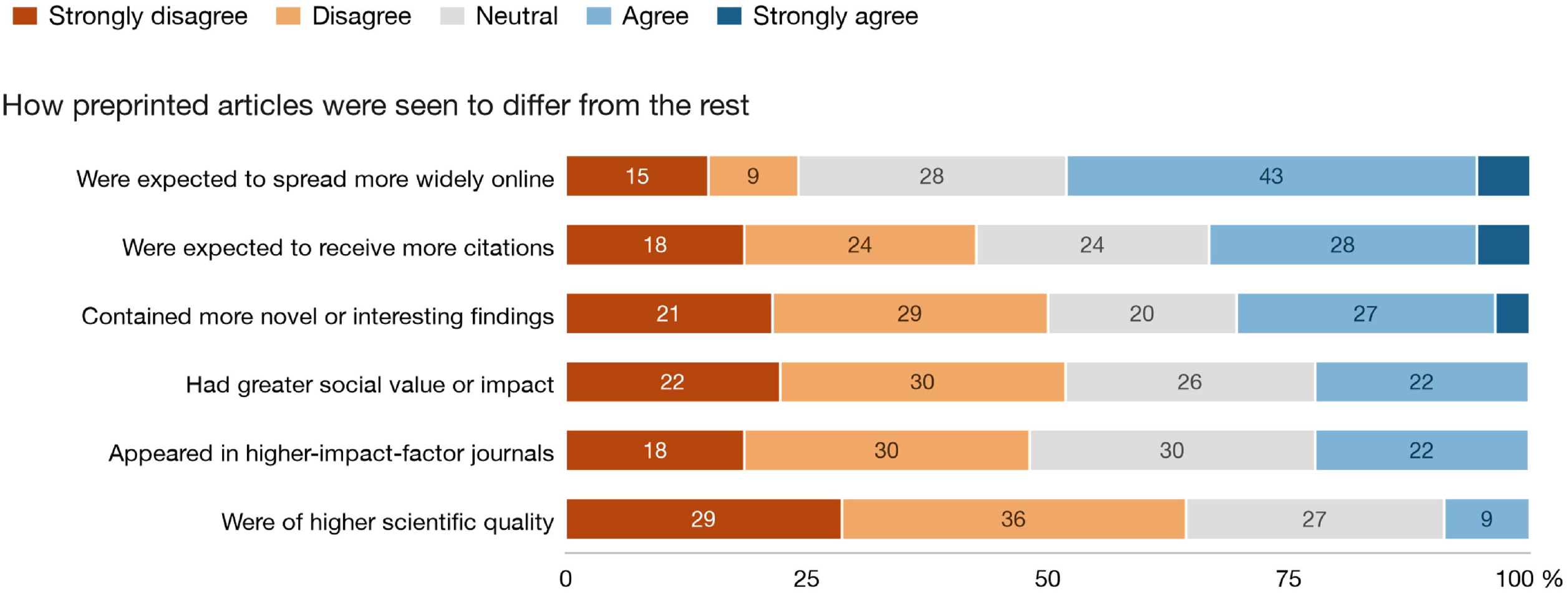
Level of agreement with statements comparing articles that were posted as preprints with those that were not, among corresponding authors who posted at least some of their articles as preprints (n = 54–56 per statement). Numbers inside the bars are percentages of respondents; segments below 7% are left unlabelled

Data on articles published by respondents in scientific journals in the last five years, in authorship positions other than corresponding author, and on the deposition of these articles as preprints are presented in the Supplementary Material.

## DISCUSSION

To our knowledge, this is the first survey on preprints adoption in a setting that combines substantial scientific output with persistent funding constraints - a highly relevant context for understanding how open science practices are adopted in resource-limited systems. Furthermore, most survey respondents had never deposited a preprint. Fraser et al. (2022) recommended targeting exactly this group in future studies, to determine whether the answers of authors who are less engaged with preprints differ from those of authors who use them.

Our results show that the low rate of adoption was due mainly to a lack of intention to deposit articles as preprints rather than to a lack of knowledge about them. Among non-depositing respondents, not wanting to deposit was the most endorsed reason (57.6%), whereas being unaware of the option at the time was endorsed by a third (35.3%) and rejected by half (50.6%). Unawareness remains a barrier for a subset of researchers, but it is not the dominant one, and the open responses point the same way: of the 22 respondents who gave an additional reason, 19 described a deliberate decision and 3 mentioned unawareness. This contrasts with earlier findings (Fraser et al., 2022), in which the strongest reason for not depositing was simply being unaware of the option at the time, leading the authors to conclude that a lack of knowledge about preprints plays the most important role. The difference shifts the focus of any intervention, because an information campaign addresses the minority reason in our sample and not the majority one. A recent international survey (Pennington et al., 2026) of 3,017 researchers across 24 disciplines supports this trend, showing that while preprints awareness reached 80%, usage remained below 50% — though in Medicine and Dentistry, adoption reached 53%. Thus, awareness of preprints is no longer the main limiting factor in the health fields, which is consistent with what our respondents report.

Our closed items did not identify why research productivity scholarship holders choose not to deposit preprints. None of the seven items on external barriers was endorsed by more than 30% of the respondents, and neutral answers were frequent, reaching 51.8% on the item about harm to the research. The one item that respondents did endorse was the statement that they did not want to deposit (57.6%), which records the decision without giving its reason. The free-text answers fill that gap. Nineteen of the 22 informative answers described a decision taken in full knowledge of the option, and the most frequent single ground was not a low estimate of the value of preprints but the view that peer review should come before public dissemination, treated as a requirement rather than as a matter of utility. Therefore, awareness was not the obstacle for these respondents; this is the finding that separates our results from those of previous surveys of authors who had already deposited preprints. Additionally, some responses revealed misconceptions regarding editorial workflows, such as fears that journals reject preprinted papers or that preprints cannot be withdrawn. Although unsupported by contemporary publishing policies, these misconceptions still act as perceived barriers.

One of the benefits of preprint deposition is related to the attention a study receives (Chiarelli et al., 2019). Studies have shown that journal articles previously posted on preprint servers received more attention, in terms of citations and Altmetric scores, than articles that were not previously posted, which points to another advantage of using preprints (Fraser et al., 2020; Fu & Hughey, 2019; Serghiou & Ioannidis, 2018). A recent study also showed a positive correlation between the number of citations to preprints and the number of citations to the peer reviewed articles (Sarkis-Onofre et al., 2023). Among respondents who had experience with preprints, however, few reported having obtained these benefits, either in terms of citations (17.3%) or in terms of online dissemination (16.9%), and on both items the modal answer was neutral (34.6% and 41.5%). Low agreement combined with this level of neutrality is better read as an absence of evidence available to the respondent than as an absence of benefit, since authors are not routinely informed of the citations or the readership of their own preprints. Another benefit of preprint deposition highlighted in the literature is the rapid dissemination of research findings, which proved particularly important during the COVID-19 pandemic. In our sample this was the leading motivation for depositing preprints, with 69.8% of respondents agreeing or strongly agreeing, and it also ranked first in Fraser et al. (2022), where 91.5% agreed.

Fraser et al. (2022) tested the self-selection bias postulate, according to which authors preferentially choose their best work to deposit as preprints and concluded that their results conflicted with it: most authors did not appear to select articles on the basis of subjective criteria, even though 43.5% expected more citations and 69.3% expected wider online sharing for the articles they had deposited. Our results point in the same direction, and more strongly. In all six items in Fig. 4, research productivity scholarship holders attributed less advantage to the articles they had deposited than the bioRxiv authors did. They rejected more firmly that these articles were of higher scientific quality (64.3% disagreement vs.43.3%), had greater social value (51.8% vs. 42.1%), contained more novel findings (50.0% vs. 39.6%) or appeared in journals with higher impact factors (48.1% vs. 44.1%), and a smaller share expected more citations (33.4% agreement, vs. 43.5%) or wider online dissemination (48.2% vs. 69.3%). These comparisons are descriptive and were not tested, and our sample of depositors is small, so the direction of the difference rather than its size is what the data support. It should also be noted that both studies measured how authors perceived the articles they had deposited, not which articles they chose to deposit, our findings provide further evidence that they did not report the pattern the postulate predicts. Since Fraser et al. (2022) recommended additional studies comparing their results with those from other servers and other research disciplines, our findings provide that comparison, in a different country and across a range of fields and demonstrate that the self-selection hypothesis fails to hold in an emerging research ecosystem.

One result, however, is specific to our setting: among the same respondents, the expectation of wider online dissemination (48.2%, Fig. 4) was almost three times the benefit they reported having obtained (16.9%, Fig. 2b), a gap of 31.3 percentage points. However, the two items are not worded identically. The first asks whether the respondent expected the articles deposited as preprints to be disseminated more widely than those not deposited, whereas the second asks whether deposition brought benefits in terms of online dissemination. Part of the gap may therefore follow from the difference in wording. The same pair of items in Fraser et al. (2022), however, produced values that were close to each other (69.3% and 66.7%), which indicates that the wording alone does not create a gap of this size. Because online sharing depends on readers who follow preprint servers in the same field and country, one reading of our result is that the network effect behind the expected gain does not materialize where the practice is uncommon. An alternative reading cannot be excluded, namely that our respondents were not in a position to observe the dissemination of their own preprints, which the 41.5% of neutral answers to that item would also be consistent with.

Finally, results from recent international survey (Pennington et al., 2026) indicate considerable levels of awareness and use among mid- and senior-career researchers, those in Medicine and Dentistry, and researchers based in the Americas - characteristics broadly consistent with our sample. This survey also highlights, in line with prior studies (Fecher et al., 2015; Houtkoop et al., 2018), that determining the factors behind open science adoption remains challenging, given the complex interactions between discipline, country, career stage and education background.

Several limitations of this study should be acknowledged. First, the response rate of 9.0% limits the generalizability of our conclusions. However, low response rates are common in email surveys of academic populations: Fraser et al. (2022) obtained fully completed questionnaires from 5.9% of a validated list of 24,633 addresses, even though those addresses had been taken from recently deposited preprints. Our rate was therefore higher than in that study, but it remains low in absolute terms, and some of the invitations may never have reached their recipients, since the addresses were collected from the Lattes Curriculum platform, from recently published articles, or from the authors’ institutional webpages, which are fewer current sources than the address declared in a recent preprint. It is also possible that the researchers we contacted were less interested in the discussion of open science practices, but our data do not allow us to distinguish between these explanations. Finally, most of the researchers who answered the survey had never deposited a preprint, which makes it harder to understand the motivations and concerns of those who have.

In terms of the context we assessed, we believe that training early career researchers on the benefits of preprint deposition would be valuable, not as a way of supplying information to researchers who already have it, but as a way of normalizing the practice among the next generation of authors, for whom the decision has not yet been settled. This follows from our results: information alone does not change an informed decision, so the case for training rests on the authors whose position is still forming rather than on those who have already declined. CAPES, through its Portal de Periódicos, has been offering training sessions on the use of open science tools, which could contribute to this process (CAPES, 2026). As for future studies, understanding the behavior of researchers in different health subject areas and of early career researchers could be an important step towards monitoring the evolution of preprint deposition. The high proportion of neutral answers to our items suggests that the reasons behind an informed decision not to deposit were not among the options we offered. Qualitative designs, such as interviews or focus groups, would therefore be better suited to identify them.

## CONCLUSION

Among the Brazilian holders of CNPq Research Productivity Grants who responded to the survey, preprint use was limited mainly by a lack of intention to deposit rather than a lack of awareness. Rapid dissemination and community feedback were the primary motivations for preprints usage, whereas a preference for formal peer review prior to public dissemination represented the principal barrier to adoption.

## Availability of data and materials

The data are available on the Open Science Framework platform (https://osf.io/futch/files/osfstorage).

## Tables Captions

**Table 1**.Characteristics of the study sample (n = 143)

