## Supplemental Material for "Motivation and concerns regarding the use of preprints among Brazilian health researchers: a survey"

| **Raw data used in the figure 1** | | | | | | |
| --- | --- | --- | --- | --- | --- | --- |
| **Considering only articles deposited as preprints, please indicate your level of agreement with the following statements.** | **Strongly disagree**  **n (%)** | **Disagree**  **n (%)** | **Neutral**  **n (%)** | **Agree**  **n (%)** | **Strongly agree**  **n (%)** | **Total**  **N** |
| The decision to deposit articles as preprints was made freely by the author(s). | 2  (3.8) | 7  (13.2) | 2  (3.8) | 27  (50.9) | 15  (28.3) | **53** |
| Depositing articles as preprints was necessary to comply with an institutional open access/preprint policy. | 17  (32.1) | 19  (35.8) | 6  (11.3) | 10  (18.9) | 1  (1.9) | **53** |
| Depositing articles as preprints was necessary to comply with an open access/preprint policy from a funding agency. | 21  (39.6) | 22  (41.5) | 7  (13.2) | 3  (5.7) | 0  (0) | **53** |
| Depositing articles as preprints was usually my suggestion. | 4  (7.5) | 18  (34) | 10  (18.9) | 12  (22.6) | 9  (17) | **53** |
| Depositing articles as preprints was usually suggested by my coauthors. | 6  (11.5) | 17  (32.7) | 14  (26.9) | 11  (21.5) | 4  (7.7) | **52** |

| **Raw data used in the figure 2A** | | | | | | |
| --- | --- | --- | --- | --- | --- | --- |
| **Considering only articles deposited as preprints, please indicate your level of agreement with the following statements.** | **Strongly disagree**  **n (%)** | **Disagree**  **n (%)** | **Neutral**  **n (%)** | **Agree**  **n (%)** | **Strongly agree**  **n (%)** | **Total**  **N** |
| Preprints were deposited to increase awareness of my/our research. | 7  (13.2) | 12  (22.6) | 10  (18.9) | 21  (39.6) | 3  (5.7) | **53** |
| Preprints were deposited to claim priority for my/our findings. | 8  (15.1) | 14  (26.4) | 8  (15.1) | 19  (35.8) | 4  (7.5) | **53** |
| Preprints were deposited to benefit the scientific enterprise. | 16  (30.2) | 16  (30.2) | 15  (28.3) | 5  (9.4) | 1  (1,9) | **53** |
| Preprints were deposited to receive more feedback on my/our work. | 5  (9.4) | 13  (24.5) | 9  (17) | 23  (43.4) | 3  (5.7) | **53** |
| Preprints were deposited to share my/our findings more rapidly. | 2  (3.8) | 9  (16.7) | 5  (9.4) | 26  (49.1) | 11  (20.7) | **53** |
| **Raw data used in the figure 2B** | | | | | | |
| **Considering only articles deposited as preprints, please indicate your level of agreement with the following statements.** | **Strongly disagree**  **n (%)** | **Disagree**  **n (%)** | **Neutral**  **n (%)** | **Agree**  **n (%)** | **Strongly agree**  **n (%)** | **Total**  **N** |
| Preprint deposition had benefits in terms of citations. | 11  (21.1) | 14  (26.9) | 18  (34.6) | 6  (11.5) | 3  (5.8) | **52** |
| Preprint deposition had benefits in terms of other forms of online dissemination, for example, sharing on social media. | 11  (20.7) | 11  (20.7) | 22  (41.5) | 4  (7.5) | 5  (9.4) | **53** |

| **Raw data used in the figure 3** | | | | | | |
| --- | --- | --- | --- | --- | --- | --- |
| **Considering only respondents who reported not depositing any articles as preprints, please indicate your level of agreement with the following statements.** | **Strongly disagree**  **n (%)** | **Disagree**  **n (%)** | **Neutral**  **n (%)** | **Agree**  **n (%)** | **Strongly agree**  **n (%)** | **Total**  **N** |
| I was unaware of the option to deposit preprints at that time. | 33  (38.8) | 10  (11.8) | 12  (14 .1) | 23  (27.1) | 7  (8.2) | **85** |
| The journal(s) to which I intended to submit did not allow prior dissemination as preprints. | 12  (14.2) | 13  (15.3) | 36  (42.3) | 16  (18.8) | 8  (9.4) | **85** |
| I was not permitted to deposit these articles as preprints, for example, due to institutional or funding agency policies. | 32  (38.5) | 16  (19.3) | 26  (31.3) | 5  (6) | 4  (4.8) | **83** |
| I did not want to deposit these articles as preprints. | 10  (11.8) | 2  (2.3) | 24  (28.2) | 16  (18.8) | 33  (38.8) | **85** |
| I wanted to deposit these articles as preprints, but my coauthors disagreed. | 51  (62.2) | 13  (15.8) | 16  (19.5) | 2  (2.4) | 0  (0) | **82** |
| Depositing a preprint would have had negative effects on my/our research. | 16  (19.3) | 10  (12) | 43  (51.8) | 10  (12) | 4  (4.8) | **83** |
| I did not want anyone else to see my/our preprint and publish before us. | 25  (29.8) | 15  (17.9) | 28  (33.3) | 10  (11.9) | 6  (7.1) | **84** |
| My our findings were controversial and needed to be evaluated through peer review before being made publicly available. | 42  (50.6) | 18  (21.7) | 18  (21.7) | 4  (4.8) | 1  (1.2) | **83** |
| My/our work did not fit the scope of any preprint repository. | 27  (32.5) | 14  (16.9) | 35  (42.2) | 4  (4.8) | 3  (3.6) | **83** |

| **Raw data used in the figure 4** | | | | | | |
| --- | --- | --- | --- | --- | --- | --- |
| **Considering only respondents who reported depositing some articles as preprints, please indicate your level of agreement with the following statements.** | **Strongly disagree**  **n (%)** | **Disagree**  **n (%)** | **Neutral**  **n (%)** | **Agree**  **n (%)** | **Strongly agree**  **n (%)** | **Total**  **N** |
| Articles deposited as preprints were of higher scientific quality than those not deposited as preprints. | 16  (28.6) | 20  (35.7) | 15  (26.8) | 5  (8.9) | 0  (0) | **56** |
| Articles deposited as preprints contained more interesting/novel findings than those not deposited as preprints. | 12  (21.4) | 16  (28.6) | 11  (19.6) | 15  (26.8) | 2  (3.6) | **56** |
| Articles deposited as preprints had greater social value/impact than those not deposited as preprints. | 12  (22.2) | 16  (29.6) | 14  (25.9) | 12  (22.2) | 0  (0) | **54** |
| Articles deposited as preprints were published in journals with higher impact factors than those not deposited as preprints. | 10  (18.5) | 16  (29.6) | 16  (29.6) | 12  (22.2) | 0  (0) | **54** |
| I expected articles deposited as preprints to receive more citations than those not deposited as preprints. | 10  (18.5) | 13  (24.1) | 13  (24.1) | 15  (27.8) | 3  (5.6) | **54** |
| I expected articles deposited as preprints to be disseminated more widely online, for example on social media, than those not deposited as preprints. | 8  (14.8) | 5  (9.3) | 15  (27.8) | 23  (42.6) | 3  (5.6) | **54** |

**Open-Ended Responses**

Question: Was there any additional reason that led you not to deposit these articles as preprints?

| **Response no.** | **Translated response** | **Codebook category** |
| --- | --- | --- |
| 1 | I prefer to believe in the peer review process as a safe tool for validating my work. | 1a. Normative objection: peer review as required validation |
| 2 | In fact, I lacked reasons to deposit the article as a preprint if it was already being submitted for publication. | 1b. No perceived added value or benefit |
| 3 | I was unaware of it, but I am also not interested in publishing before finalizing the manuscripts. | 1a. Normative objection: peer review as required validation \| secondary: 2a. Unaware of the option or did not seek information |
| 4 | I am not convinced that it is a good thing. | 1a. Normative objection: peer review as required validation |
| 5 | 1) I have the impression that there are many poor-quality articles deposited as preprints, especially from the COVID period. 2) I observed that, after months in the repository, many articles received few or no views and virtually no comments. The original idea from the field of physics, that there would be many comments similar to peer review, has not been confirmed; in fact, even traditional journals are currently unable to find reviewers. From a practical point of view, depositing in a preprint repository seems useless. | 1b. No perceived added value or benefit \| secondary: 1a. Normative objection: peer review as required validation |
| 6 | Scientifically, it seems to be a questionable practice. | 1a. Normative objection: peer review as required validation |
| 7 | There was no reason for not having a preprint. | X. Unclassifiable response |
| 8 | I do not agree with the publication of preprints. | 1a. Normative objection: peer review as required validation |
| 9 | Totally against it. | 1a. Normative objection: peer review as required validation |
| 10 | I think it is partly the work involved in making a new submission, combined with the lack of urgency in publication. | 1e. Procedural burden and lack of habit \| secondary: 1b. No perceived added value or benefit |
| 11 | Previous unfavorable experience - depositing in a preprint repository created an opportunity for predatory journals to contact me requesting publication. After it was published, I was unable to remove it from the preprint server. The policy was very poor and unfavorable, and I no longer considered publishing in this way. Also because I consider peer review an essential step before giving visibility to a publication. | 1c. Perceived risk to formal publication \| secondary: 1a. Normative objection: peer review as required validation |
| 12 | I did not look for information about preprints in the journals. | 2a. Unaware of the option or did not seek information |
| 13 | The manuscript goes to preprint, and then if the journal does not accept it, I cannot publish it anywhere. | 1c. Perceived risk to formal publication |
| 14 | No interest. | 1b. No perceived added value or benefit |
| 15 | Simply lack of knowledge. | 2a. Unaware of the option or did not seek information |
| 16 | I do not agree. | 1a. Normative objection: peer review as required validation |
| 17 | Perhaps lack of habit, lack of knowledge about why to do it, and additional work. | 1e. Procedural burden and lack of habit \| secondary: 2a. Unaware of the option or did not seek information |
| 18 | It recently happened to us that a thesis published in an institutional repository was cited by the leading journal in the field as a reason for not accepting the article for publication, indicating that the article had already been published online. They agreed to publish it after we removed it from the institutional repository. Therefore, we have avoided preprints. | 1c. Perceived risk to formal publication |
| 19 | I do not have a formed opinion about the impact of preprints. | 2b. No formed opinion about preprints |
| 20 | It did not count on the curriculum. | 1d. Absence of career or evaluation incentive |
| 21 | I did not think it was necessary. | 1b. No perceived added value or benefit |
| 22 | I am totally against the idea of preprints. | 1a. Normative objection: peer review as required validation |
| 23 | I do not see any benefit in depositing preprints. | 1b. No perceived added value or benefit |

**Coding frame.** Theme 1, informed rejection (n = 19): 1a peer review as required validation; 1b no perceived added value; 1c perceived risk to formal publication; 1d absence of career or evaluation incentive; 1e procedural burden and lack of habit. Theme 2, non-informed non-use (n = 3): 2a unaware of the option or did not seek information; 2b no formed opinion about preprints. X, unclassifiable (n = 1). Four responses state two distinct reasons and carry a secondary code; the primary code alone is used for the counts, so that no response is counted twice. Counts describe the respondents who answered this optional open field and are not estimates of prevalence in the sample.

**Supplementary Table – Preprints Survey**

**Regarding articles that you published in a scientific journal in any position other than corresponding author in the last 5 years, select which statement applies:**

| **Statement** | **%** | **n** |
| --- | --- | --- |
| All these articles were also deposited as preprints. | 0.68% | 1 |
| Some of these articles were also deposited as preprints. | 40.41% | 59 |
| None of these articles were deposited as preprints. | 58.90% | 86 |

**You indicated that you deposited preprints for some of the articles you published in a scientific journal in any position other than corresponding author in the last 5 years. Considering these articles, indicate your level of agreement with the following statements: (Please choose the appropriate response for each item)**

| **Statement** | **Strongly disagree (%)** | **Strongly disagree (n)** | **Disagree (%)** | **Disagree (n)** | **Neutral (%)** | **Neutral (n)** | **Agree (%)** | **Agree (n)** | **Strongly agree (%)** | **Strongly agree (n)** | **Total** |
| --- | --- | --- | --- | --- | --- | --- | --- | --- | --- | --- | --- |
| Articles deposited as preprints were of higher scientific quality than those not deposited as preprints. | 27.27% | 15 | 41.82% | 23 | 18.18% | 10 | 10.91% | 6 | 1.82% | 1 | 55 |
| Articles deposited as preprints contained more interesting/novel findings than those not deposited as preprints. | 25.93% | 14 | 38.89% | 21 | 16.67% | 9 | 16.67% | 9 | 1.85% | 1 | 54 |
| Articles deposited as preprints had greater social value/impact than those not deposited as preprints. | 24.07% | 13 | 44.44% | 24 | 20.37% | 11 | 11.11% | 6 | 0 | 0 | 54 |
| Articles deposited as preprints were published in journals with higher impact factors than those not deposited as preprints. | 24.07% | 13 | 31.48% | 17 | 27.78% | 15 | 16.67% | 9 | 0 | 0 | 54 |
| I expected articles deposited as preprints to receive more citations than those not deposited as preprints. | 18.52% | 10 | 22.22% | 12 | 33.33% | 18 | 22.22% | 12 | 3.70% | 2 | 54 |
| I expected articles deposited as preprints to be disseminated more widely online (e.g., on social media) than those not deposited as preprints. | 18.52% | 10 | 14.81% | 8 | 24.07% | 13 | 37.04% | 20 | 5.56% | 3 | 54 |

**Considering only articles that were deposited as preprints, indicate your level of agreement with the following statements: (Please choose the appropriate response for each item).**

| **Statement** | **Strongly disagree (%)** | **Strongly disagree (n)** | **Disagree (%)** | **Disagree (n)** | **Neutral (%)** | **Neutral (n)** | **Agree (%)** | **Agree (n)** | **Strongly agree (%)** | **Strongly agree (n)** | **Total** |
| --- | --- | --- | --- | --- | --- | --- | --- | --- | --- | --- | --- |
| The decision to deposit articles as preprints was the free decision of the author(s). | 1.89% | 1 | 7.55% | 4 | 13.21% | 7 | 58.49% | 31 | 18.87% | 10 | 53 |
| It was necessary to deposit articles as preprints to comply with an institutional open access/preprint policy. | 32.69% | 17 | 32.69% | 17 | 19.23% | 10 | 13.46% | 7 | 1.92% | 1 | 52 |
| It was necessary to deposit articles as preprints to comply with a funding agency's open access/preprint policy. | 32.69% | 17 | 40.38% | 21 | 17.31% | 9 | 9.62% | 5 | 0 | 0 | 52 |
| It was usually my suggestion to deposit articles as preprints. | 24.53% | 13 | 30.19% | 16 | 22.64% | 12 | 16.98% | 9 | 5.66% | 3 | 53 |
| It was usually my co-authors' suggestion to deposit articles as preprints. | 11.54% | 6 | 19.23% | 10 | 26.92% | 14 | 32.69% | 17 | 9.62% | 5 | 52 |
| Preprints were deposited to increase awareness of my/our research. | 15.09% | 8 | 16.98% | 9 | 33.96% | 18 | 30.19% | 16 | 3.77% | 2 | 53 |
| Preprints were deposited to claim priority for my/our findings. | 15.09% | 8 | 26.42% | 14 | 22.64% | 12 | 30.19% | 16 | 5.66% | 3 | 53 |
| Preprints were deposited to benefit the scientific enterprise (a science-based project developed by, or in cooperation with, the private sector). | 26.42% | 14 | 35.85% | 19 | 30.19% | 16 | 5.66% | 3 | 1.89% | 1 | 53 |
| Preprints were deposited to receive more feedback on my/our work. | 18.52% | 10 | 20.37% | 11 | 14.81% | 8 | 38.89% | 21 | 7.41% | 4 | 54 |
| Preprints were deposited to share my/our findings more rapidly. | 11.11% | 6 | 12.96% | 7 | 20.37% | 11 | 40.74% | 22 | 14.81% | 8 | 54 |
| Preprint deposition had benefits in terms of citations. | 18.52% | 10 | 27.78% | 15 | 37.04% | 20 | 11.11% | 6 | 5.56% | 3 | 54 |
| Preprint deposition had benefits in terms of other forms of online dissemination (e.g., sharing on social media). | 16.98% | 9 | 24.53% | 13 | 41.51% | 22 | 9.43% | 5 | 7.55% | 4 | 53 |

**You indicated that you deposited preprints for all articles that you published in a scientific journal in any position other than corresponding author in the last 5 years. Considering these articles, indicate your level of agreement with the following statements: (Please choose the appropriate response for each item).**

| **Statement** | **Strongly disagree (%)** | **Strongly disagree (n)** | **Disagree (%)** | **Disagree (n)** | **Neutral (%)** | **Neutral (n)** | **Agree (%)** | **Agree (n)** | **Strongly agree (%)** | **Strongly agree (n)** | **Total** |
| --- | --- | --- | --- | --- | --- | --- | --- | --- | --- | --- | --- |
| The decision to deposit articles as preprints was the free decision of the author(s). | 0 | 0 | 0 | 0 | 1 | 1 | 0 | 0 | 0 | 0 | 1 |
| It was necessary to deposit articles as preprints to comply with an institutional open access/preprint policy. | 0 | 0 | 0 | 0 | 1 | 1 | 0 | 0 | 0 | 0 | 1 |
| It was necessary to deposit articles as preprints to comply with a funding agency's open access/preprint policy. | 0 | 0 | 0 | 0 | 1 | 1 | 0 | 0 | 0 | 0 | 1 |
| It was usually my suggestion to deposit articles as preprints. | 0 | 0 | 0 | 0 | 1 | 1 | 0 | 0 | 0 | 0 | 1 |
| It was usually my co-authors' suggestion to deposit articles as preprints. | 0 | 0 | 0 | 0 | 1 | 1 | 0 | 0 | 0 | 0 | 1 |
| Preprints were deposited to increase awareness of my/our research. | 0 | 0 | 0 | 0 | 1 | 1 | 0 | 0 | 0 | 0 | 1 |
| Preprints were deposited to claim priority for my/our findings. | 0 | 0 | 0 | 0 | 1 | 1 | 0 | 0 | 0 | 0 | 1 |
| Preprints were deposited to benefit the scientific enterprise (a science-based project developed by, or in cooperation with, the private sector). | 0 | 0 | 0 | 0 | 1 | 1 | 0 | 0 | 0 | 0 | 1 |
| Preprints were deposited to receive more feedback on my/our work. | 0 | 0 | 0 | 0 | 1 | 1 | 0 | 0 | 0 | 0 | 1 |
| Preprints were deposited to share my/our findings more rapidly. | 0 | 0 | 0 | 0 | 1 | 1 | 0 | 0 | 0 | 0 | 1 |
| Preprint deposition had benefits in terms of citations. | 0 | 0 | 0 | 0 | 1 | 1 | 0 | 0 | 0 | 0 | 1 |
| Preprint deposition had benefits in terms of other forms of online dissemination (e.g., sharing on social media). | 0 | 0 | 0 | 0 | 1 | 1 | 0 | 0 | 0 | 0 | 1 |

**You indicated that you did not deposit preprints for any of the articles that you published in a scientific journal in any position other than corresponding author in the last 5 years. Considering these articles, indicate your level of agreement with the following statements: (Please choose the appropriate response for each item)**

| **Statement** | **Strongly disagree (%)** | **Strongly disagree (n)** | **Disagree (%)** | **Disagree (n)** | **Neutral (%)** | **Neutral (n)** | **Agree (%)** | **Agree (n)** | **Strongly agree (%)** | **Strongly agree (n)** | **Total** |
| --- | --- | --- | --- | --- | --- | --- | --- | --- | --- | --- | --- |
| I was unaware of the option to deposit preprints at that time. | 35.37% | 29 | 17.07% | 14 | 15.85% | 13 | 24.39% | 20 | 7.32% | 6 | 82 |
| The journal(s) to which I wanted to submit did not allow prior dissemination of preprints. | 18.52% | 15 | 11.11% | 9 | 45.68% | 37 | 14.81% | 12 | 9.88% | 8 | 81 |
| I was not permitted to deposit these articles as preprints (e.g., due to institutional or funding agency policies). | 42.50% | 34 | 16.25% | 13 | 32.50% | 26 | 5.00% | 4 | 3.75% | 3 | 80 |
| I did not want to deposit these articles as preprints. | 10.98% | 9 | 6.10% | 5 | 34.15% | 28 | 14.63% | 12 | 34.15% | 28 | 82 |
| I wanted to deposit these articles as preprints, but my co-authors disagreed. | 53.09% | 43 | 14.81% | 12 | 30.86% | 25 | 0 | 0 | 1.23% | 1 | 81 |
| Depositing a preprint would have had negative effects on my/our research. | 32.10% | 26 | 14.81% | 12 | 39.51% | 32 | 9.88% | 8 | 3.70% | 3 | 81 |
| I did not want anyone else to see my/our preprint and publish before us. | 29.63% | 24 | 16.05% | 13 | 29.63% | 24 | 17.28% | 14 | 7.41% | 6 | 81 |
| My/our findings were controversial and needed to be assessed by peer review before being made public. | 41.25% | 33 | 22.50% | 18 | 30.00% | 24 | 5.00% | 4 | 1.25% | 1 | 80 |
| My/our work did not fit the scope of any preprint repository. | 31.25% | 25 | 20.00% | 16 | 41.25% | 33 | 6.25% | 5 | 1.25% | 1 | 80 |
